# Calibrated simulations for dynamic focusing of ultrasound through the temporal window

**DOI:** 10.64898/2026.01.27.26344890

**Authors:** Ehsan Dadgar-Kiani, Vibha Hebbale, Grace Attalla, John L Alvarez, Suraya Dunsford, Kevin A Caulfield, Cameron H. Good, Andrew D Krystal, Leo P Sugrue, Joline M Fan, Elsa Fouragnan, Samuel Pichardo, Kim Butts Pauly, Keith R Murphy

**Affiliations:** Attune Neurosciences, San Francisco, CA, USA; Brain Research and Imaging Centre, Faculty of Health, University of Plymouth, Plymouth, UK; School of Psychology, Faculty of Health, University of Plymouth, Plymouth, UK; Department of Psychiatry, Medical University of South Carolina, Charleston, SC, USA; Department of Psychiatry, University of California San Francisco, San Francisco CA; Department of Radiology and Biomedical Imaging, University of California San Francisco, San Francisco CA; Department of Neurology, University of California San Francisco, San Francisco CA; Radiology, Cumming School of Medicine, University of Calgary; Hotchkiss Brain Institute, University of Calgary; Department of Radiology, Stanford University, Stanford, CA, USA

**Keywords:** neuromodulation, focused ultrasound, deep brain stimulation, safety, acoustic simulation

## Abstract

Focused ultrasound can be delivered through the temporal window to modulate heterogeneously located brain areas. Acoustic simulations allow for safety assessments when dynamically targeting brain structures, but the mismatch between simulation and measured focal pressure can vary across the steerable range due to mechanically inaccurate assumptions made about the skull and transducer. Here, we describe efficient methods for simulation-measurement calibration using axisymmetric projections and sparse sampling across a 3D steerable subspace encompassing deep brain targets across 157 subjects. To address the simulation-reality mismatch in skull transmission, we used the measured and predicted pressure values through eight human temporal window fragments to derive an optimized bone attenuation coefficient. Collectively, the calibration framework and optimized temporal window coefficients can be used broadly across studies to improve the accuracy of reporting and dependent safety assessment for personalized neuromodulation treatments.

## MAIN

Ultrasound neuromodulation is a rapidly emerging brain stimulation technology being actively researched and developed for therapeutic clinical applications^1–4^ and causal functional brain investigation^5–9^. As ultrasonic modulatory technology develops, a key consideration is how simulations of pressure and temperature rise can inform the safety and accuracy of beam steering capabilities of multi-element transducers^10,11^. Researchers often use transducers with a pre-defined and often fixed focal position, allowing for a single calibration value between the transducer drive voltage and focal pressure in free-field conditions^10^. In contrast, multi-element arrays in a two-dimensional (2D) plane allow for dynamic steering of an ultrasound focus across three-dimensional (3D) space. With large mechanical or electrical cross-talk, there may be substantial errors in simulation unless differential calibration is used when steering to different locations in 3D space^12–15^. If the location of the array is known relative to the subject’s anatomy, an approach of calibration for every point in space can be used to target heterogeneous brain space in a highly precise manner (**Fig. 1a**). While this capability extends researchers’ placement and targeting capabilities, highly sampled voltage-pressure maps across a given steering range may be impractical, particularly if each independent ultrasound system requires its own calibration. Although it is possible to characterize fields based on exact relative coordinates derived from individual modeling, logistical challenges arise when the device is deployed in the field and target position varies across each use. To address these challenges as the use of focused ultrasound neuromodulation scales, we examined the relationship between spatial calibration resolution and accuracy within a single plane and in 3D space.

**Figure 1.**
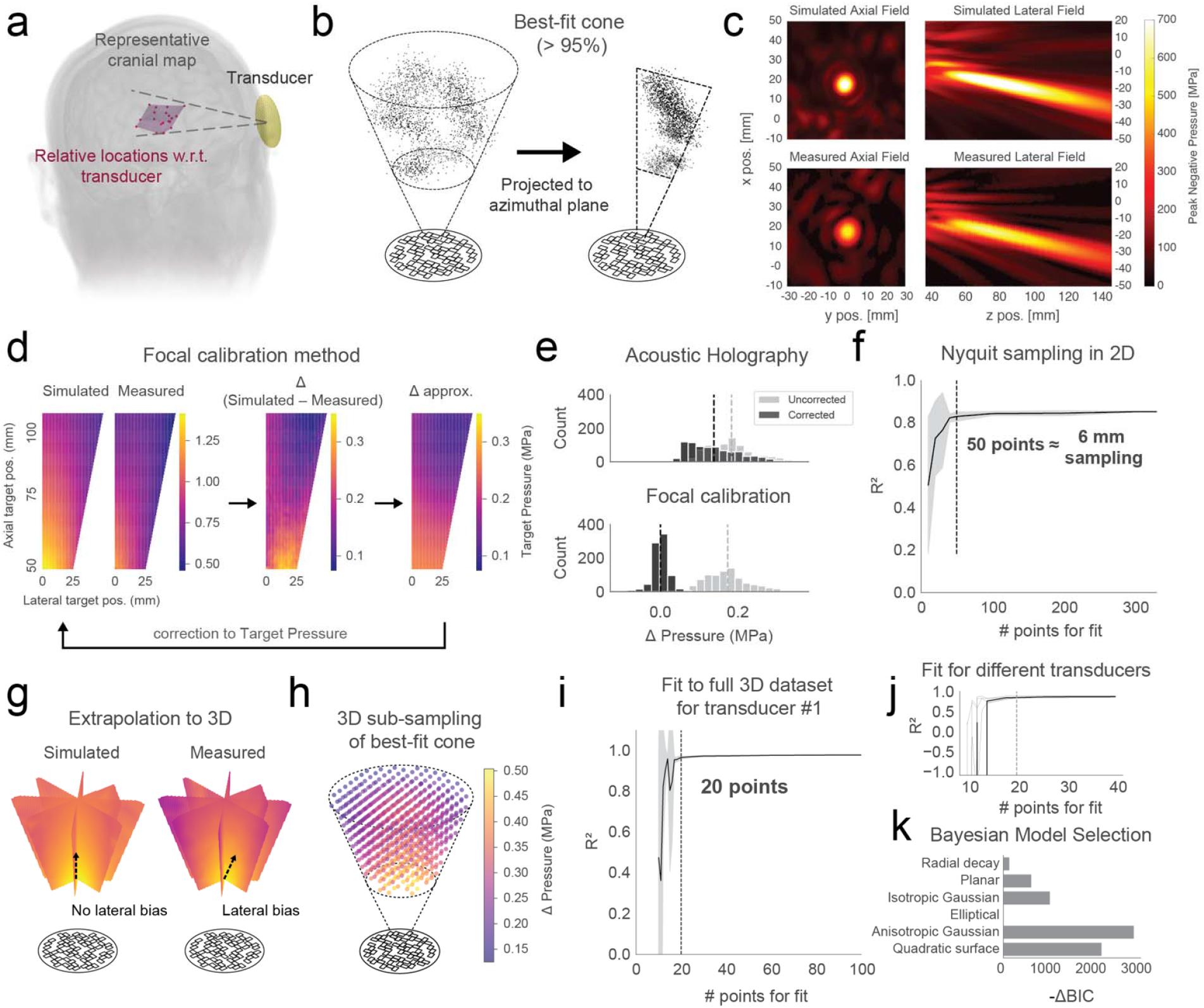
3D steering simulation calibration for FUS neuromodulation. **a**) Target coordinates relative to a single transducer array across heterogeneous individual anatomy. **b**) Target coordinates in 3D space (left) can be flattened (right) to represent axial and lateral distances within an azimuthal calibration plane. **c**) Comparisons between axial and lateral planes of simulated and measured ultrasound pressure fields. **d**) A schematic of the focal correction method. Difference between simulated and measured focal pressure maps at Nyquist-sampling across the calibration plane shows a low frequency gradient mismatch, which can be approximated by a global fit and used to correct a simulation’s focal pressure while steering within the calibration plane. Compared to acoustic holography, the focal calibration method better minimizes the differences between simulated and measured focal pressure. **e**) Within the Nyquist-sampled calibration plane, anisotropic Gaussian function fit strength related to equidistant number of points sampled for fitting the mismatch (Δ) between 2D focal pressure maps from simulation and measurement. **f**) Planar measurements of focal pressure at different azimuthal angles shows nonuniformity not evident in simulated planes. Arrow indicates the lateral spatial bias observed in measurements. **g**) Visualization of 3D calibration field fully sampled at 6 mm is well fit by **h**) a 3D anisotropic gaussian function. **i**) Similar fits with comparable convergence patterns around 20 points were obtained for other transducers (N = 5). **j**) Bayesian model selection can be used to identify the appropriate model type that captures the simulation-reality mismatch without unnecessary complexity.

## RESULTS

We first determined the spatial extent of focused ultrasound (FUS) targeting by extracting the relative target coordinates from four bilateral deep brain targets; the caudate, amygdala, centromedian nucleus of the thalamus, and subthalamic nucleus across 157 subjects. Since the 2D array coverage has a circular aggregate symmetry, the coordinates were then flattened into a single plane, capturing the axial and lateral steering depth by zeroing the azimuthal angle (**Fig. 1b**). The calibration space was then defined as the truncated cone that captured 95% of the target coordinates in the axial and lateral domain. In principle, this statistical approach to defining calibration space can be applied to any study where a set of brain target coordinates is known relative to a transducer, which may narrow or broaden calibration space. Initially we examined whether acoustic holography could be used to create source pressure patterns sufficient for creating accurate simulation of pressure outputs with our array. As described previously^15^, the phase and amplitude for each array element was independently collected at a depth of 30 mm from the transducer radiating surface. The plane was then backpropagated to derive a source pressure mapping for each element (**Supplemental Fig. 1a**). Using the reconstructed pressure sources for each element yielded significant mismatch between measurement and simulation peak pressures across the steerable range in excess of 20% (**Supplemental Fig. 1b**). This mismatch is likely due to large crosstalk of elements that is not accounted for when measuring element activity in isolation^14,16^. Thus, while acoustic holography may be sufficient for dynamically steered focal calibration in arrays with highly isolated elements, partially isolated bulk piezoelectric arrays may require direct measure across the steering space.

To directly build a mapping between simulation and measurement we captured the peak focal pressure in simulation and measured field when steering to fixed locations across the steering range. The peak was chosen rather than intended focal position even in the event that the two were slightly offset. This ensures the calibration is conservatively safe by assuming the plane’s maximum pressure. To identify a minimal and practical sampling frequency needed to characterize a 2D calibration space, the steering range was initially sampled at the Nyquist spatial frequency of 1.5 mm based on the free-field wavelength of ∼3 mm at 500 kHz. For all positions, the hydrophone scan was set to sample the focal plane at 6 points per wavelength, below Nyquist frequency. The simulated focal pressure was then compared to the experimentally measured focal pressure, determined by taking the peak within the focal target’s axial plane (**Fig. 1c**). The focal pressure obtained when steering the array to all coordinates in the calibration space defines a focal pressure mapping, and can be constructed from both simulated and experimental focal pressures (**Fig. 1d**). The difference between these two maps defines the discrepancy between simulated and measured target pressures, and contained a gradual, low frequency component with the near-field lateral target points showing the largest discrepancy. The data were well captured with an anisotropic Gaussian function fit (**Fig. 1f**). Using random step-wise subsampling of target pressure discrepancies across the calibration space, we found that 50 points produced a strong fit (*R*^2^ = 0.83) with negligible improvements from increased sampling (**Fig. 1f**). Fifty points correspond to an equidistant spacing of 6 mm between sampled points within the calibration space. Going forward, we utilized an equidistant 6-mm spacing of target points when calibrating 2D planes and 3D cones. Also, given that we observed similar distributions of spatial frequencies in simulated focal pressure maps across concentric circle, Fermat spiral, and normally distributed element grid designs (**Supplemental Fig. 1a, b**), we expect that this simulation-reality correction for focal pressure maps can be applied to a multitude of array designs.

In theory, arrays with axial symmetry may also contain symmetric cross-talk and calibration fields, and different coordinates with conserved depth-angle along that axis should produce highly similar pressure profiles. In this case, a single plane could theoretically be measured as a proxy for planes rotated along the orthogonal axis. As expected, acoustic simulations produced largely axisymmetric pressure fields for both axisymmetric and asymmetric arrays. However, modest differences were found when experimentally measuring 8 planes’ focal pressure maps at 45° azimuthal steps for our random non-axisymmetric array, potentially due to manufacturing variability or crosstalk not captured by the simulations (**Fig. 1g, Supplemental Fig. 3**). In the event that these planes are largely conserved, or that the relationship between them is well characterized, a 2D plane may be rotated around the axis of symmetry to produce a calibration value for points in 3D space. However, in the event of plane mismatch, a 3D characterization is necessary for each transducer. To identify a minimum viable number of points, we sampled the entire 3D steerable range at 6 mm resolution (**Fig. 1h**). Based on elbow detection of *R*^*2*^ fit, we found that the 3D space was well fit with just 20 points (**Fig. 1i**), likely due to the low spatial frequencies composing the discrepancy between simulation and measurement. This approach also generalized to five other transducers with the same element configurations (**Fig. 1j, Supplemental Fig. 2a**), with each showing similar convergence patterns of fit at just about 20 points (**Supplemental Fig. 2b, c)**. Using Bayesian model selection, other functions used to fit the data underperformed relative to the 3D anisotropic Gaussian function (**Fig. 1k**). Thus, this calibration sampling may be used with other array designs operated at 500 kHz, which may be validated with at least one high-resolution field sampling. While elbow detection balances accuracy with efficiency and diminishing returns from adding more points, other objective methods for selecting a sampling rate may also be used, such as a mean discrepancy threshold.

The calibration method described can be applied in simulations to approximate the output of ultrasound neuromodulation devices in water, which closely approximate the acoustic properties of the brain. As both water and brain tissue are highly transmissive of ultrasound, pressure measurements in water can generally be assumed to represent the maximal pressure in the brain, assuming that the skull is acoustically transparent and the absence of standing waves from the skull interior^15,16^. In contrast, the change in acoustic properties as waves interact with the skull can induce appreciable phase shifting, absorption, reflection, and scattering, substantially altering ultrasound transmission. Lower resolution computed tomography (CT) or magnetic resonance imaging (MRI) of gross bone structure can allow for coarse estimation of these features. However, cortical bone contains finer structural elements that are not practically observable with imaging^17^. In particular, the trabecular bone has numerous heterogeneous cavities with rapid changes in acoustic impedance, which can result in substantial scattering. Indeed, earlier works demonstrated that the majority of ultrasound attenuative losses across calvarial bone are from scattering rather than absorption^1^. Given the practical resolution limits of current imaging and simulation frameworks, microstructure scattering cannot be properly simulated and attenuation by scattering is heavily underestimated. A workaround to this issue is the addition of a loss term to acoustic simulation space assigned specifically to bone. Within pseudospectral time-domain simulations, as implemented formally by k-Wave^18,19^, attenuation coefficients are modeled with a frequency power-law α(f)=α □f^y^ together with the matched dispersion term implemented via fractional-Laplacian operators. Using a viscoelastic full-wave model as implemented formally by BabelBrain^20^, attenuation and dispersion are modeled by viscoelastic constitutive laws with relaxation or Q-factor terms, assigning longitudinal and shear losses per voxel from CT-derived property maps. While these terms are effective at representing loss, accurate assignment accounting for bone loss has not yet been determined.

To optimize and understand the attenuation associated with dynamic steering through the temporal window, we first approximated the device position on each of 5 cadaver skulls. We temporarily placed 5 mm silicone pads between the device and bone to mimic the soft tissues of the scalp which are absent on the skulls. Using a guide mock transducer, we measured the distance from mock transducer fiducials to the temporal window bone and marked the relative orthogonal projection point. The mock transducer was then fitted with posts to replicate the depth to skull while a surrounding bracket was mounted to the skull (**Fig. 2a**). Both left and right temporal windows were then cut from the skull, degassed in water, and scanned with a zero echo time (ZTE) MRI sequence as previously described^21^; this process was performed for each of 5 skulls, yielding 8 different skull fragments after 2 fragments were discarded due to cracking damage along the sutures (**Fig. 2a**). To examine whether our fragments were representative of live subjects’ scanned temporal windows, we used each subject’s ZTE scan and the transducer elements’ orthogonal projections to virtually extract both the left and right temporal windows from 157 subjects’ MRI scans (**Fig. 2b**). The ex-vivo fragments closely resembled the virtual temporal windows in both skull thickness and incidence angle as measured through each transducer element’s projection (**Fig. 2c**). This indicates that findings and simulations performed with the bone fragments would be applicable to live subjects.

**Figure 2.**
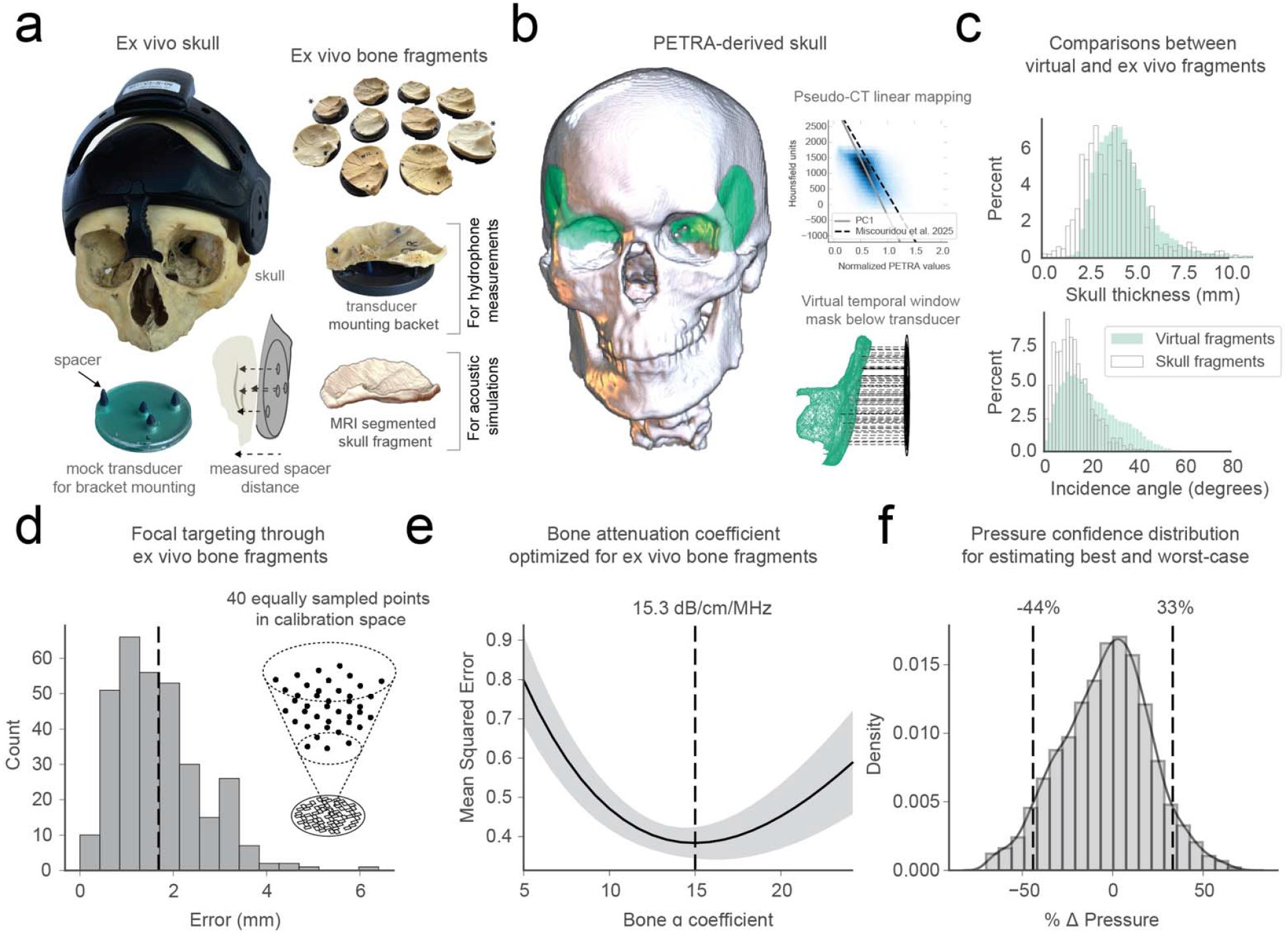
Optimization of simulation attenuation with heterogeneous temporal window transmit measurement. **a**) Wearable device is placed on each human skull sample for measurement of distance from transducer control points to temporal window. A mock transducer with those recorded distances set with spacers is used to further mount a bracket for holding the ultrasound transducer in a fixed location relative to the skull. The bracket and transducer location are captured in the MRI space of each skull fragment. Image of mounted temporal window skull fragments. Asterisks indicate two fragments that were omitted due to damage. **b**) Normal projections from each transducer element to ZTE-derived skull masks were used to extract “virtual” temporal windows. An example virtual temporal window is shown. CT and ZTE scans of five bone fragments were also used to derive a linear relationship between Hounsfield units and ZTE voxel units, in close agreement with mappings in previous literature. **c**) Population distribution of virtual temporal windows extracted from 157 subjects are compared to simulated projections from ex vivo skulls for element angle of incidence and temporal window thickness. **d**) Focal targeting was performed at forty equally-sampled points in the calibration space for each ex vivo bone fragment, and the histogram depicts the spatial targeting error (mm). **e**) Plot of mean squared error between measurement and simulation focal pressure with stepwise homogenous assignment of ultrasound wave absorption in bone. **f**) At the optimal attenuation coefficient (□ = 15.3 dB/cm/MHz), the 95th percentile over-estimation of focal pressure corresponds to 33%.

For each fragment, element phase delays were calculated through acoustic simulation time reversal targeting each of 40 locations equally sampled across the calibration space (**Fig. 2d, Supplemental Fig. 4b,c**). The mean targeting error across each combination of 8 bone fragments and 40 spatial targets was found to be 1.68 ± 0.96 mm off from the intended target (**Fig. 2d**). Irrespective of the targeting error, we extracted the peak pressure from the target plane to ensure conservative safety calibrations. We then compared these measured pressures with simulated peak pressures obtained from forward acoustic simulations as we incrementally modified the homogenous bone attenuation coefficient (α □). The mean square error between simulation and reality was minimized with a value of 15.3 dB/cm/MHz (**Fig. 2e**), in close alignment with previous experimental estimations of skull bone attenuation of 13.3 dB/cm and 16.6 dB/cm at 1 MHz^17,18^. To derive conservative boundaries when using this optimized value, we calculated the 5^th^ and 95^th^ percentile pressure error across simulation-measurements. The 5^th^ percentile overestimation and 95^th^ percentile underestimation found that the pressure could be 44% lower or 33% higher, respectively, than the simulated value (**Fig. 2f**).

## DISCUSSION

In this study we provide a methodology for calibrating simulation of ultrasound arrays foci across a steering range. We further optimize simulation attenuation to improve simulation accuracy and to understand simulation error boundaries with respect to peak pressures. This will ultimately improve ultrasound neuromodulation reporting and our understanding of safe stimulation parameters. Devices that lack steering capability or have a preset number of focal positions may have simpler calibration requirements, such as making a single measurement per device. Furthermore, a device with measured axial symmetry may require less measurements. While non-definitive, we provide a suggestive algorithm for calibration steps depending on the transducer type (i.e., single vs. multi-element) (**Supplemental Fig. 5**). For skull attenuation, we optimize for accuracy and include a distribution of estimates to reflect the heterogeneity in bone microstructure that impacts ultrasound transmission. This approach improves upon our earlier practices, which reported single, conservative estimates that consider the skull as highly transmissive^2^. In this way, variant methods can be purposed toward capturing more realistic values rather than conservatively high values. Nevertheless, methods of attenuation use should be clearly specified in the methodology^10^ to properly guide replication efforts and safety^2^. In addition to the data-driven approach to calibration field size creation used here, other viable methods may include population level biometric data based on head anatomy^19^, or large MRI datasets^20–22^. For instance, bizygomatic breadth, or the maximum width measured between the outer edges of the cheekbones, has been captured for large sample populations using calipers which offer 5^th^ percentile and 95^th^ percentile ranges for both sexes^23^. Furthermore, field size may be determined by an acceptable ratio between the main lobe and side or grating lobes present^10^.

Regardless of the methodology used for calibration, discrepancies between the intended and measured focal position may arise from subtle mechanical registration error, or element phase distortion associated with device cross-talk (**Fig. 1c**). Thus, capturing the actual peak rather than taking a direct measurement at the intended focus is strongly recommended to ensure that the calibration conservatively accounts for the largest possible biomechanical effects. This approach enables the accurate assessment of safety limits derived from peak pressures such as Mechanical Index (MI) or spatial peak pulse average intensity (ISPPA). In some cases, the focus may contain secondary fields likely associated with transducer element mechanical cross-talk stemming from limited physical isolation. This cross-talk may shift the peak pressure closer to the array face than intended, despite conservation of steering angle. If there is only one stimulation focus, then the axial plane may be used to determine the appropriate lateral plane for capturing the calibration focal peak. In the event that multiple foci are targeted to a cross-beam location arising from multiple transducers, the peak plane will be at the intended axial target plane (**Supplemental Fig. 6**) at the superposition of the two beams, as implemented by numerous teams^2,24,25^. In these instances, measurement of the peak at the intended focal plane may be advisable to avoid reporting overestimated peak pressures. Regardless of configuration, the measured field volume should be sufficiently large to capture the focal peak, where the size should be expanded until the field maximum value is not found at the edge of the measurement window. Furthermore, the expected pressure from the calibration must be accounted for within the source pressure assignment, or within a multi-stage error assessment that also includes biological inputs to the system, such as the skull.

Although this work emphasizes estimation of pressure and mechanical index, thermal derivatives are also important for ultrasound neuromodulation safety^11^. Previous works have found absorption to account for ∼16.8% of attenuative losses^26^. Thus, thermal simulations may be performed where the calibrated pressure field is derived and passed into a thermal simulation, such as Pennes’ bioheat equation simulation. Here, the absorption coefficient can be set to 16.8% of the optimized attenuation value, as the remaining fraction likely represents scattering. Collectively, these methodologies will improve the accuracy of simulations and thus, the assumptions of safety when using steerable ultrasound arrays for human applications.

## METHODS

### Ultrasound transducer and drive system

A custom sparse 2D array containing 64 elements (**Supplemental Fig. 2a**) was manufactured (Sonic Concepts Inc.) and driven at 500 kHz by a custom drive system, ATTN201 (Attune Neurosciences Inc.). Each array element was rectangular (4.15 mm x 2.7 mm). When performing scan tank measurements, the array was pulsed at 70 V (peak-to-peak), a 5 Hz pulse repetition frequency, and a duty cycle of 1%.

When simulating variant ultrasound array designs (**Supplemental Fig. 2a**), a concentric circle layout with 3.8 mm diameter circular elements, a Fermat spiral layout with 3.8 mm diameter circular elements, and a grid with 3.38 mm square elements were used. All variant designs contained 64 elements.

### Hydrophone scan measurements

Hydrophone scan tank measurements were made using a capsule hydrophone (ONDA HGL-400) calibrated at 500 kHz mounted in the ONDA AIMS III scan tank (Product # AST3-L-3). Directivity loss compensation was calculated using a Bessel function fit as specified by the manufacturer. Water was degassed and filtered continued with an Aquas-10 Water Conditioning system (Onda product # AQUAS-10-110V).

When steering the ultrasound transducer to a given target point, the hydrophone and scan tank system were used to measure the peak pressure at each point in an XY plane (0.5 mm resolution) at the target point and orthogonal to the transducer array. The peak pressure value in this plane was used as the focal pressure. Directivity correction was performed to each pressure measurement using the hydrophone’s provided Bessel function, which considers the angle between the transducer and hydrophone to yield a pressure correction factor.

### Calibration model

To fit the calibration model to the measured spatial discrepancy between measured and simulated peak pressures, we optimized the following generalized Gaussian function:

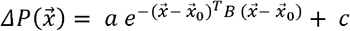

The scalar constants *a, c*, and matrix *B* were set by minimizing the mean squared error between measured and simulated peak pressures for all target points and skull fragments. For 2D spatial calibrations (**Fig. 1d-f**), *B* was represented as a 2×2 diagonal matrix, whereas for the 3D spatial calibrations (**Fig. 1g-k**), *B* was represented as a 3×3 diagonal matrix. All optimizations and analyses were performed using the Python language and SciPy package.

### Skull mounting

Cadaver skulls were obtained from Skulls Unlimited International, Inc. Transducer positioning was set by placing an ATTN201 headset on the skull with 5 mm foam padding at the skull intersection points to mimic scalp thickness offset. The distance between each fiducial and the orthogonal skull intercept was measured by first inserting a thin metal rod through ⅛ inch drilled holes in each fiducial until it touched the skull. A 3D printed washer was fitted tightly to the rod and pressed flush against the outer surface of the transducer while inserting the rod to ensure orthogonal alignment. To mark the length of the measurement rod from the fiducial to the skull, the rod and tightly fit washer were removed from the headset placed on the skull and reinserted to the corresponding fiducial on an identical headset until the washer was flush against the outer surface. A felt marker was then used to mark the distance from the tip of the rod to the inner transducer surface, then measured with calipers once the rod was removed. The intersect position on the skull was then marked by dabbing the measurement rod in red acrylic paint and re-inserting the rod into the headset on the skull until the washer was flush with the outer transducer surface and the rod was therefore touching the skull. This process was repeated for each fiducial.

To secure the bracket to the skull at the correct distance and angle from each fiducial, conical shaped spacers the length of the previously measured distances between the fiducials and orthogonal skull were 3D printed and super glued to a fiducial puck at each location (**Fig. 2a**). A sharpie was used to mark the center of the fiducial and assist in aligning the spacer with the center of each fiducial. This puck was then placed in the bracket and the fiducial spacers were aligned with the corresponding paint marks on the skull. To secure the bracket to the skull, holes were first drilled into the bracket. Clay was fit around the skull to allow for orthogonal drilling through the bracket in 3 locations. The skull segment surrounding the bracket was traced then extracted with a drill. Finally, the bracket was secured to the skull fragment by aligning the fiducial puck with the paint marks on the skull segment once again then adding screws to hold the bracket. Glue was added sparingly to secure the screws in place then the fiducial puck was removed. Prior to experiments, skulls were degassed in a plexiglass chamber filled with deionized water using a vacuum pump at -30 Hg for at least 48 hours. In total, 10 skull fragments from 5 cadaver skulls were obtained. Two fragments and their corresponding MRI scans were not used for acoustic simulations due to damaged bone.

### MRI Scanning

Whole head scanning was performed on a Siemens MAGNETOM Cima.X 3T scanner with a 64-channel head coil. MPRAGE (0.94 mm isotropic resolution) and ZTE (0.75 mm isotropic resolution) scans were collected for each of 157 subjects while simultaneously wearing an MRI-compatible version of the mock headset, using key scan parameters derived previously^21^. ZTE scans were also acquired for 5 mounted ex vivo skull fragments with MRI fiducials while submerged in a plastic bag containing previously degassed filtered water.

### MRI Analysis

Spherical fiducial markers (8 total; 4 on each side) in the headset were visible with high MR contrast and used to align the MPRAGE and ZTE scans to a common coordinate system. Brains were extracted from the MPRAGE volumes using a custom 3D U-Net model^28^ trained on a large open source dataset^24–26^. Brains were then aligned with both the Allen and MNI reference atlases using the ANTs software package to derive target coordinates corresponding to several deep brain regions. The deep brain regions (Centromedian nucleus of the thalamus, Caudate, Amygdala, and Subthalamic Nucleus) were selected based on earlier studies using our custom ultrasound device^1,29^. Skull masks were automatically extracted from ZTE scans using another custom 3D U-Net model^28^ trained on a subset of manually annotated data.

### Acoustic Simulations

Acoustic simulations were performed in MATLAB R2024a using the k-Wave v1.4.0 toolbox and kspaceFirstOrder3D function^27^. Simulation parameters include a Courant-Friedrichs-Lewy (CFL) value of 0.3, BonA nonlinearity parameter value of 6, power law absorption exponent of 1.5, isotropic spatial sampling of 0.7 mm, and 20 cycle sinusoidal pulsing to reach steady state. Peak simulation values were captured from the entire simulation time domain. All simulations were performed on a desktop running Ubuntu 24.04.2 LTS with an NVIDIA GeForce RTX 4070 GPU.

Five rigidly aligned CT and ZTE bone fragment scans (**Supplemental Fig. 4a**) were used to derive a linear relationship between Hounsfield units and ZTE voxel units. A robust principal component analysis of corresponding CT and ZTE voxel values yielded the following equation based on the primary principal component, CT = -3609.8 × ZTE + 3258.8 (**Fig. 2b**). This fit generally agrees with that from Miscouridou et al. (2025), CT = -2929.6 × ZTE + 3274.9. While direct conversion for each fragment would have given idealized skull properties, as it was important to base our analysis on generalizable, real-world use cases, we only used this data to validate the existing ZTE-CT mapping provided in literature^28^. Using the mapping from Miscouridou et al. (2025), voxel-level ZTE values were converted to a pseudo-CT before being used to calculate speed of sound and density values for use in the acoustic wave propagation equations.

To steer through the bone fragment and maximize pressure at a given coordinate, a time reversal simulation was used to calculate the individual element phases to be used in a subsequent forward simulation. The globally optimal bone attenuation coefficient was selected that minimized the mean squared error between simulated target pressure and measured target pressure across all bone fragments and target positions.

To obtain a globally optimal bone attenuation coefficient across the 8 ex vivo temporal window bone fragments, the attenuation coefficient was first swept through a range of values from 5.0 to 25.0. For a given alpha value, we simulated the propagation of acoustic waves from transducer elements through each ex vivo bone fragment and targeting each of the forty projected coordinates (**Fig. 2d**).

### Acoustic Holography

Acoustic holography was used to calibrate a custom sparse 2D phased array transducer by measuring the complex acoustic field generated by each element over a two-dimensional plane. A capsule hydrophone (ONDA HGL-400), calibrated at 500 kHz and mounted in an ONDA AIMS III scan tank (AST3-L-3), was raster-scanned at a fixed axial depth of 30 mm across a 120 mm × 120 mm region using a uniform spatial step size of 0.7 mm. At each (x, y) position, individual array elements were driven sequentially by a Verasonics Vantage NXT system, which emitted a 500-kHz, 100-µs pulse for each element followed by a 100-µs inter-element delay. For every transmitted pulse, the resulting hydrophone waveform was recorded using a PicoScope PS5000A oscilloscope.

Recorded signals were converted from ADC units to millivolts and processed offline. Waveforms were bandpass-filtered (400–500 kHz) using a zero-phase Butterworth filter, and the analytic signal was computed via the Hilbert transform to extract instantaneous amplitude and phase. Because elements were fired with fixed timing, each element’s contribution appeared in a known temporal window. For each element at each spatial location, phase was sampled at a fixed time corresponding to the onset of the pulse, and amplitude was quantified as the peak negative pressure within a defined analysis window, using the hydrophone sensitivity of 6.2 V/MPa. These measurements were compiled into three-dimensional arrays indexed by lateral position, elevational position, and element number, yielding a complete holographic map of the element-wise acoustic field for calibration and field reconstruction.

For each element’s complex hologram, angular spectrum backpropagation was performed using the angularSpectrumCW function in the k-Wave v1.4.0 toolbox in order to recover the phase and amplitude in the source plane. For use in focusing in a forward propagation simulation, each complex source plane could be phase offset based on the element’s distance to the target, and all planes could then be summed together. For forward propagation, the same angularSpectrumCW function was used.

## Data Availability

All data produced in the present study are available upon reasonable request to the authors.

---

**Supplemental Figure 1.**
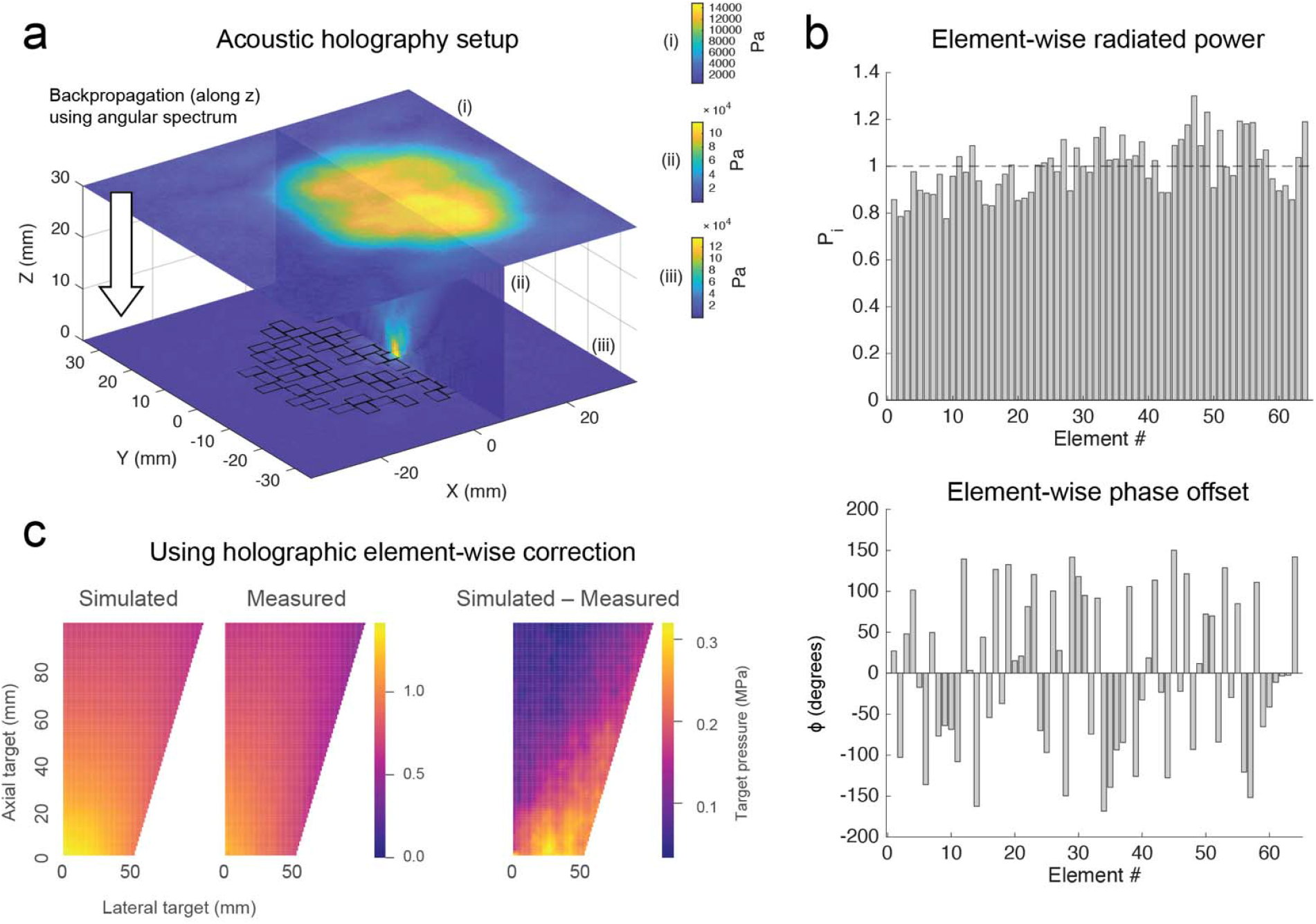
Holography-based transducer calibration is unable to capture spatially dependent cross-talk when steering a multi-element phased array. **a**) Depiction of the acoustic holography setup used to calibrate a 2D phased array transducer by measuring the complex acoustic field generated by each element over a two-dimensional plane at a distance of 30 mm from the array plane. This measured complex plane (i) from a single source element is backpropagated using the angular spectrum method to recover the source complex plane (iii). An orthogonal plane (ii) depicts the spatial convergence of the pressure field to the original source element through backpropagation. In (i), (ii), and (iii), only complex magnitude is depicted. **b**) When using the source complex planes to simulate focusing across the steering range, the discrepancy between simulated peak pressures and measured peak pressures still shows a significant spatial dependence, indicating that purely holographic-based transducer calibration is unable to fully capture the cross-talk.

**Supplemental Figure 2.**
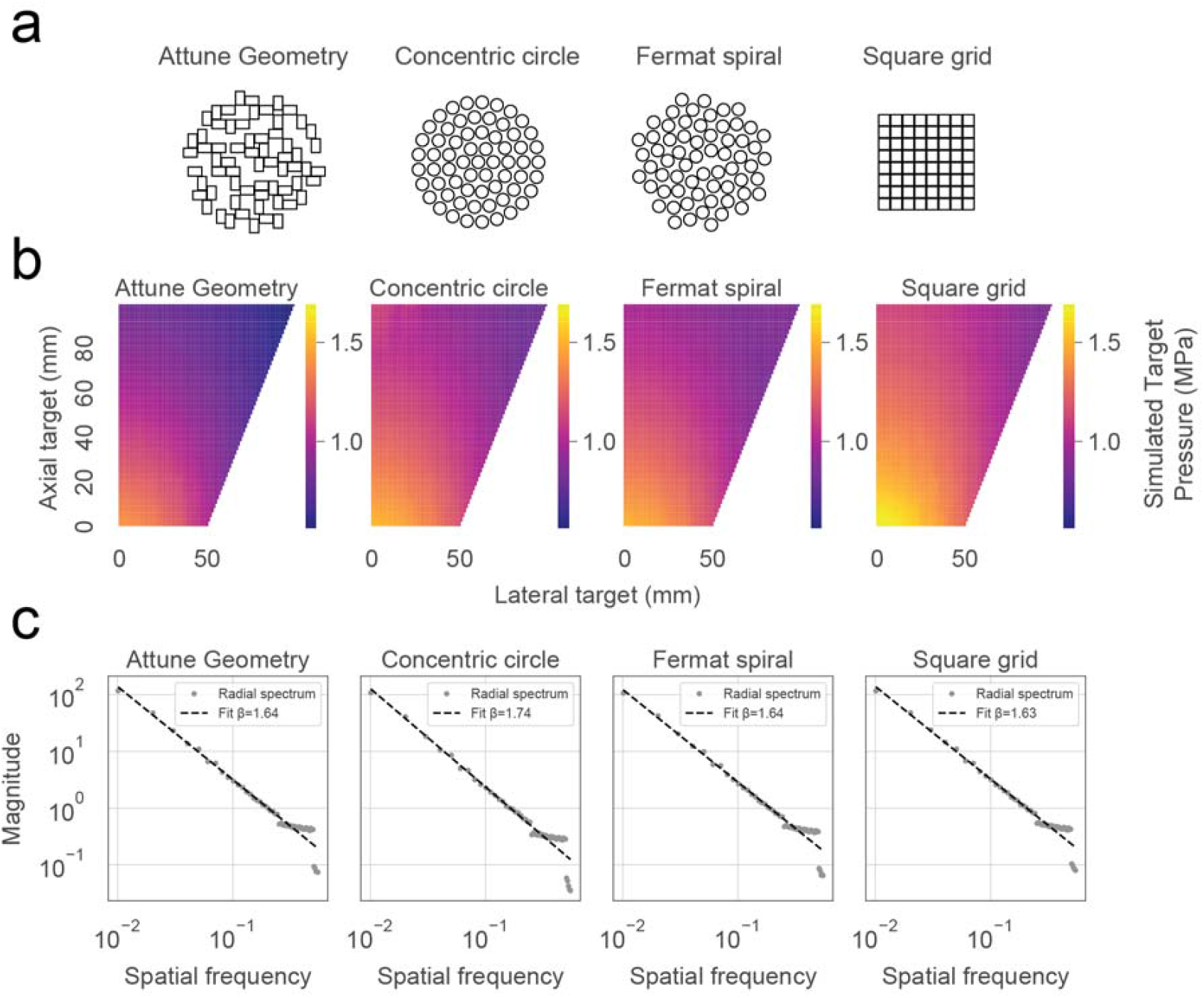
Variant ultrasound array designs tested for focal pressure output symmetry. **a**) Depictions of 2D transducer element configurations used to generate **b**) simulated focal pressure maps when steering across a 2D calibration range. **c**) All array configurations’ focal pressure maps showed comparable magnitudes across the relevant spatial frequency range.

**Supplemental Figure 3.**
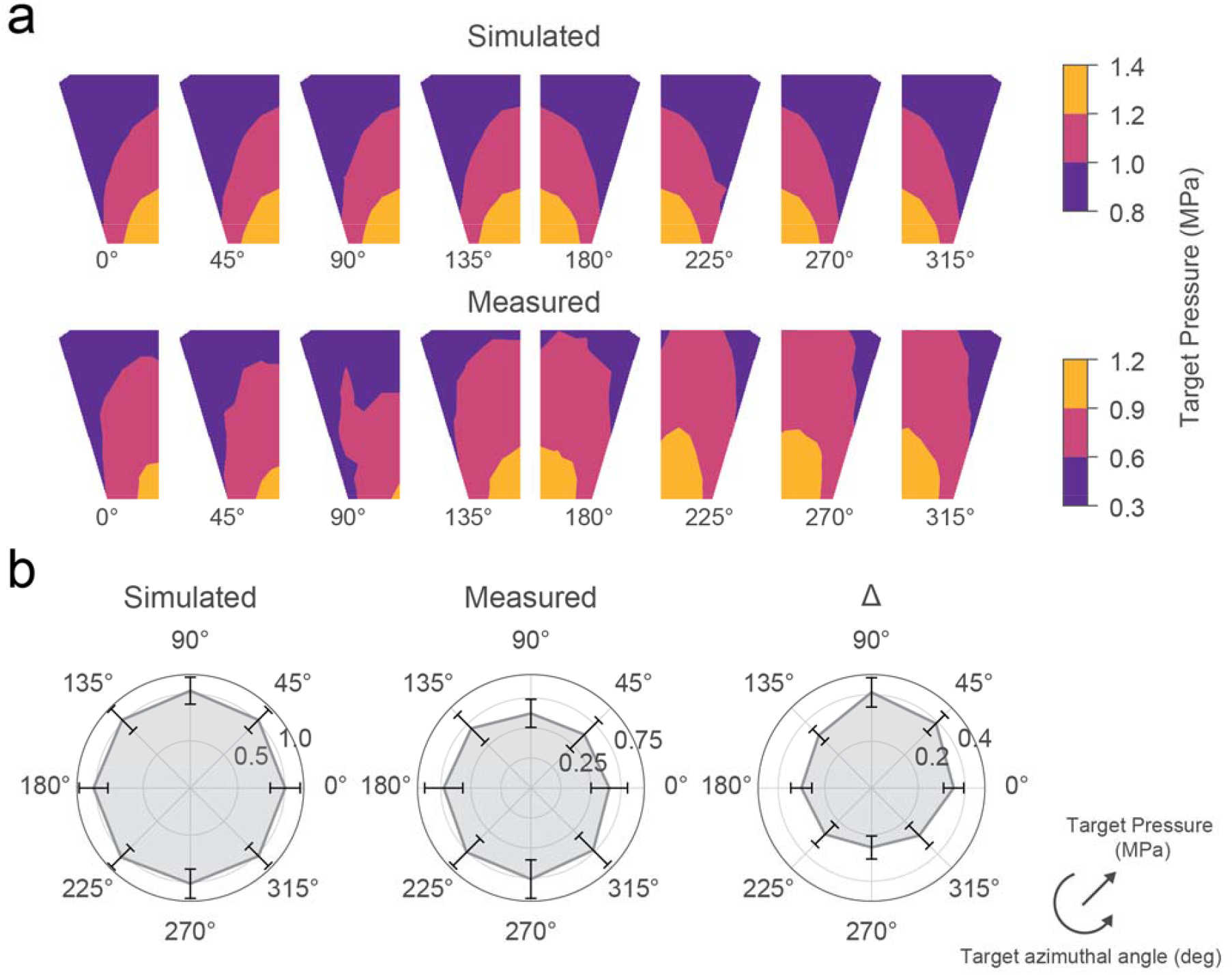
Focal pressures at variable azimuthal angles show spatial nonuniformity and lateral bias between measurement and simulation. **a)** Comparisons between simulated and measured focal pressure cross sections ranging from 0º to 315º in 45º increments. Maps were segmented into three uniform color bins to better depict the uniformity evident in simulation but absent in measurement. **b)** Radar charts of the focal pressures and discrepancies between measured and simulated values shows a lateral bias towards 45º and 90º that would not be captured in uncorrected simulations.

**Supplemental Figure 4.**
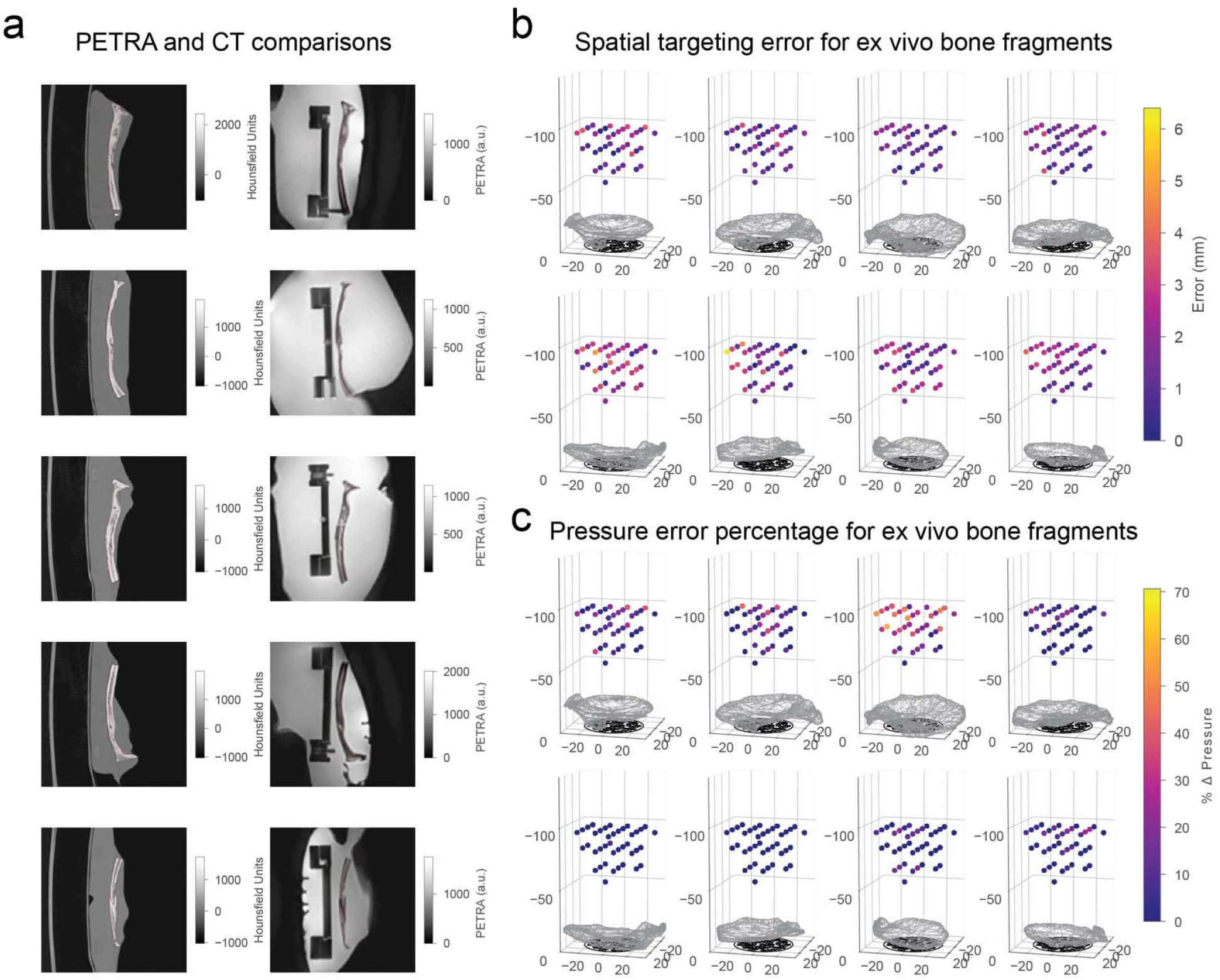
Spatial targeting errors and simulated focal pressure errors through mounted temporal window skull fragments. **a**) Comparisons of CT and ZTE scans acquired for five fragments after rigid alignment between their bone masks. **b**) Depictions of each fragment with targets used for optimizing the bone alpha coefficient. Target points are color-coded by spatial error (mm) between intended and measured focal peak in the target plane. **c**) Similar depictions of the percentage error between measured and simulated target pressure at each point when using the optimized attenuation coefficient (15.3 dB/cm/MHz).

**Supplemental Figure 5.**
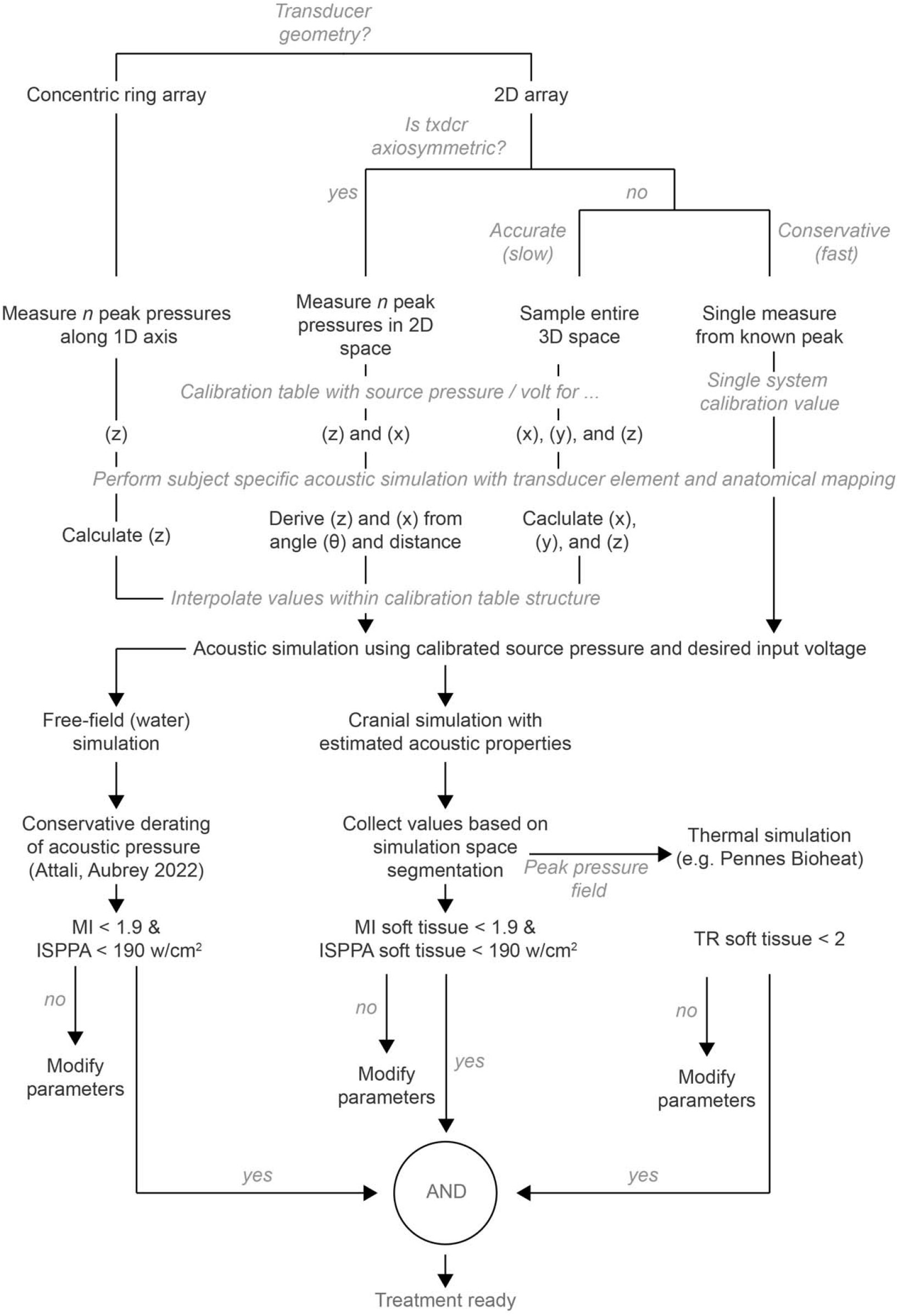
Algorithmic approach to array calibration

**Supplemental Figure 6.**
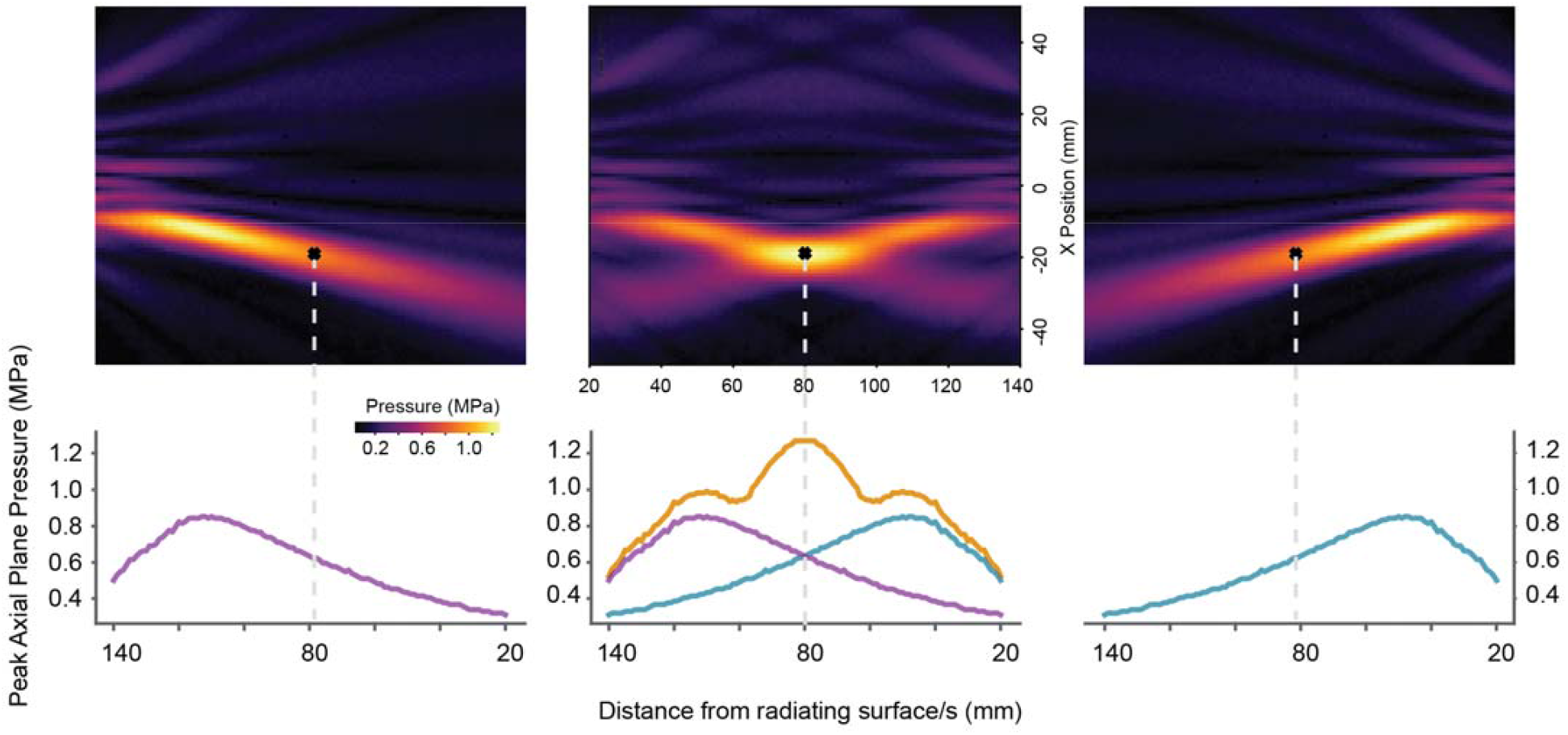
Single array focusing may result in near-field shifted peak pressure (Left, Right). Intersectional cross beams where the intended focal positions are summed exceeds the peak pressure of independent near field pressures.

